# Potential uses of AI for perioperative nursing handoffs: a qualitative study

**DOI:** 10.1101/2022.01.08.22268939

**Authors:** Christopher Ryan King, Ayanna Shambe, Joanna Abraham

## Abstract

**Objective:** Situational awareness and anticipatory guidance for nurses receiving a patient after surgery are key to patient safety. Little work has defined the role of artificial intelligence (AI) to support these functions during nursing handoff communication or patient assessment. We used interviews and direct observations to better understand how AI could work in this context.

**Materials and Methods:** 58 handoffs were observed of patients entering and leaving the post-anesthesia care unit at a single center. 11 nurses participated in semi-structured interviews. Mixed inductive-deductive thematic analysis extracted major themes and subthemes around roles for AI supporting postoperative nursing.

**Results:** Four themes emerged from the interviews: (1) Nurse understanding of patient condition guides care decisions, (2) Handoffs are important to nurse situational awareness; problem focus and information transfer may be improved by AI, (3) AI may augment nurse care decision making and team communication, (4) User experience and information overload are likely barriers to using AI. Key subthemes included that AI-identified problems would be discussed at handoff and team communications, that AI-estimated elevated risks would trigger patient re-evaluation, and that AI-identified important data may be a valuable addition to nursing assessment.

**Discussion and Conclusion:** Most research on postoperative handoff communication relies on structured checklists. Our results suggest that properly designed AI tools might facilitate postoperative handoff communication for nurses by identifying elevated risks faced by a specific patient, triggering discussion on those topics.

**LAY SUMMARY:** Nurses caring for patients after surgery make many decisions about what complications to look for and how to treat issues that arise. They rely on handoffs from prior providers to understand the patient’s background, events, and plans so far. We observed nurse handoffs after surgery where operating room nurses transfer information and their care responsibility to postoperative unit nurses to ensure care continuity. We also interviewed nurses to ask if and how artificial intelligence (AI) might help them focus their handoff communication on likely problems and generally understand the patient. Our participants stated that if AI identified likely issues, they would discuss those topics in handoff, communicate about those problems with physicians, and modify their monitoring and treatment to the level of risk faced by the patient. This finding runs against most research on improving communication, which focuses on checklists of topics to discuss. Most uses of AI for nurses focus on making specific to-do recommendations and documentation reminders and search, but we find that nurses would benefit from AI which focuses more on their understanding of the patient’s condition. These findings have major implications for the application of AI support for nurses.

## BACKGROUND AND SIGNIFICANCE

Inpatient handoffs are the transfer of responsibility, information, and control between clinical providers or teams. Incomplete or inaccurate handoffs are a source of subsequent medical errors and patient injury, [1, 2] particularly for patients undergoing major surgery.[3–5] We focus on handoffs for postoperative nursing during the operating room (OR) to post-anesthesia care unit (PACU) and PACU to ward transition for inpatient surgical populations. Handoffs are important for receiving nurses to understand the patient’s situation because residual sedation, pain, delirium, fatigue, and surgical injuries can make patient-nurse communication difficult. Additionally, the patient’s context is subject to change; old concerns may be eliminated by surgery and potential complications emerge. The data surrounding surgical patients is voluminous and diverse while simultaneously incomplete, which strains the ability of receiving nurses to review and assimilate it de novo.[6–8] Two functions of handoff are of special interest to us: situational awareness and anticipatory guidance. Situational awareness is the combination of perceiving critical factors in the environment, understanding what those factors mean for the provider’s goals, and understanding what will happen next.[9] Anticipatory guidance is the communication of likely patient status changes and plans for how to address them.[10, 11] Both of these concepts have been integrated into major quality improvement projects.[12–14] Protocolized or checklist based handoffs are employed to ensure that key information is transmitted throughout healthcare.[15–17] Similar methods of standardization have been applied to nurse-to-nurse handoffs [12, 18] and perioperative nursing handoffs specifically.[19, 20] Nevertheless, handoff-related information gaps are common for postoperative patients.[8, 21–24]

The electronic health record (EHR) has promise for mitigating and reducing these information gaps. EHRs place data at the fingertips of all providers. In theory, this ought to allow a nurse to prepare for handoff and recover from an incomplete handoff. Despite this promise, most handoff-EHR integration work does not focus on the critical functions of situational awareness and anticipatory guidance.[25] Staggers and others [26] found that existing EHR handoff summaries were too rigid and incomplete to be useful; additionally, they interfered with the receiving nurse’s encoding of information via note taking. They subsequently found nursing EHR handoff support to be poorly utilized due to these limitations.[27] One exciting development has been the use of artificial intelligence (AI) to supplement the manual review and assimilation of EHR data. For example, use of information dashboards simplify the stream of real-time data (for example, vital sign and laboratory data).[26–33] More complex contemporary systems include identification of “relevant” patient data and prediction of multiple adverse events.[32–34]

A parallel literature explores the role of AI-based prediction of adverse events of specific relevance to nursing tasks. For example, identification of patients with high risk of pressure ulcers [35, 36] or falls [37] can trigger clinical decision support for nursing interventions. The related clinical decision support literature focuses on recommending specific actions, medication dosing, and documentation reminders. [31] Only a handful of studies have considered combining these two threads with AI-based prediction to augment handoff communication. In the neonatal ICU context, Hunter and colleagues [33] used natural-language generation to summarize EHR data and generate potential problems and care plans to facilitate handoffs. Forbes and colleagues [32, 38] designed a shift-report summary for nurses including key data, diagnoses, and predicted adverse events. Hunter and Forbes’s work [32, 33, 38] suggests a new role for AI-based prediction: facilitating problem-based report and assessment during handoffs. Although the assessment of the patient’s condition is a key part of all structured handoffs, AI identification of likely complications and important data could prompt review of the most pertinent issues while being much more flexible than traditional checklist-based protocols. We previously explored related ideas at the OR to intensive care unit handoff (which often has a brief nurse-to-nurse component due to the multi-disciplinary handoff).[39] Key findings of that study were the difficulty of making EHR information universally accessible, the need to focus on AI with direct relevance to patient care, and general acceptance of blending AI risk prediction with current summaries of patient data into a handoff tool. However, the ICU shift-change and OR-ICU handoffs which have been previously studied are quite different from the OR-PACU- ward transition.

## OBJECTIVES

Before implementing AI support for postoperative handoffs, we identified key unanswered questions in prior research literature about postoperative bedside nurses as givers or receivers of handoff: (1) would postoperative nurses accept AI recommendations for handoff topics? (2) would nurses find AI-based predictions useful and relevant? (3) would nurses share a preferred presentation for AI-based predictions (making them widely accessible)? The goal of this single center qualitative study was to explore these topics with a focus on potential gaps in the postoperative nursing handoff process and how AI might fit into the situational awareness, assessment, monitoring, and communication goals of post-anesthesia care unit (PACU) and post-operative ward nurses. We intend these findings to guide subsequent design and implementation efforts.

## MATERIALS AND METHODS

### Setting

Barnes-Jewish Hospital is a 1400 bed academic medical center in St Louis, Missouri where approximately 20,000 surgeries are performed annually. We focused on the Acute and Critical Care Surgery (ACCS) division, which performs approximately 1600 inpatient surgeries annually, primarily trauma and acute abdominal surgery. All postoperative patients (other than those directly admitted to intensive care) recover from anesthesia in the PACU, a 30-bed area. Four hospital units subsequently care for ACCS patients: two dedicated hospital wards and two high-dependency units which are shared with otolaryngology, abdominal organ transplant, and hepatobiliary services.

Surgical cases were selected for direct observation from the OR schedule based on the primary surgery service (ACCS). Patients likely to be admitted to high dependency units based on their procedures were also included. Interviews and direct observations were conducted under Washington University IRB approval (#201812137 and #202009066) with the consent of the PACU nurse to shadow their activities. Written consents were changed to verbal consents with electronic provision of study information during the COVID-19 pandemic.

### Participants

Participants included the OR circulator nurse, anesthesia provider, surgery provider, PACU nurse, and wards nurse caring for these patients. Concurrently, a convenience sample of PACU and ward nurses was recruited from the above units (ACCS floor and high-dependency wards). Interviews were conducted until topic saturation.

### Description of Perioperative Handoff Processes and Care Teams

The handoff process is illustrated in Figure 1. Prior to surgery, a preoperative holding area nurse completes a health status inventory in the Epic EHR and on a paper record (Appendix S1) passed to PACU. An informal handoff is completed between the preoperative nurse and OR circulating nurse. After surgery, the patient is transported to the PACU by a surgery resident or fellow, the OR circulating nurse, and an anesthesia provider (nurse anesthetist or anesthesiology resident). OR to PACU handoff follows a protocol (Appendix S1) where the circulating nurse, surgeon, and anesthesia provider each give handoff to the PACU nurse.

**Figure 1:**
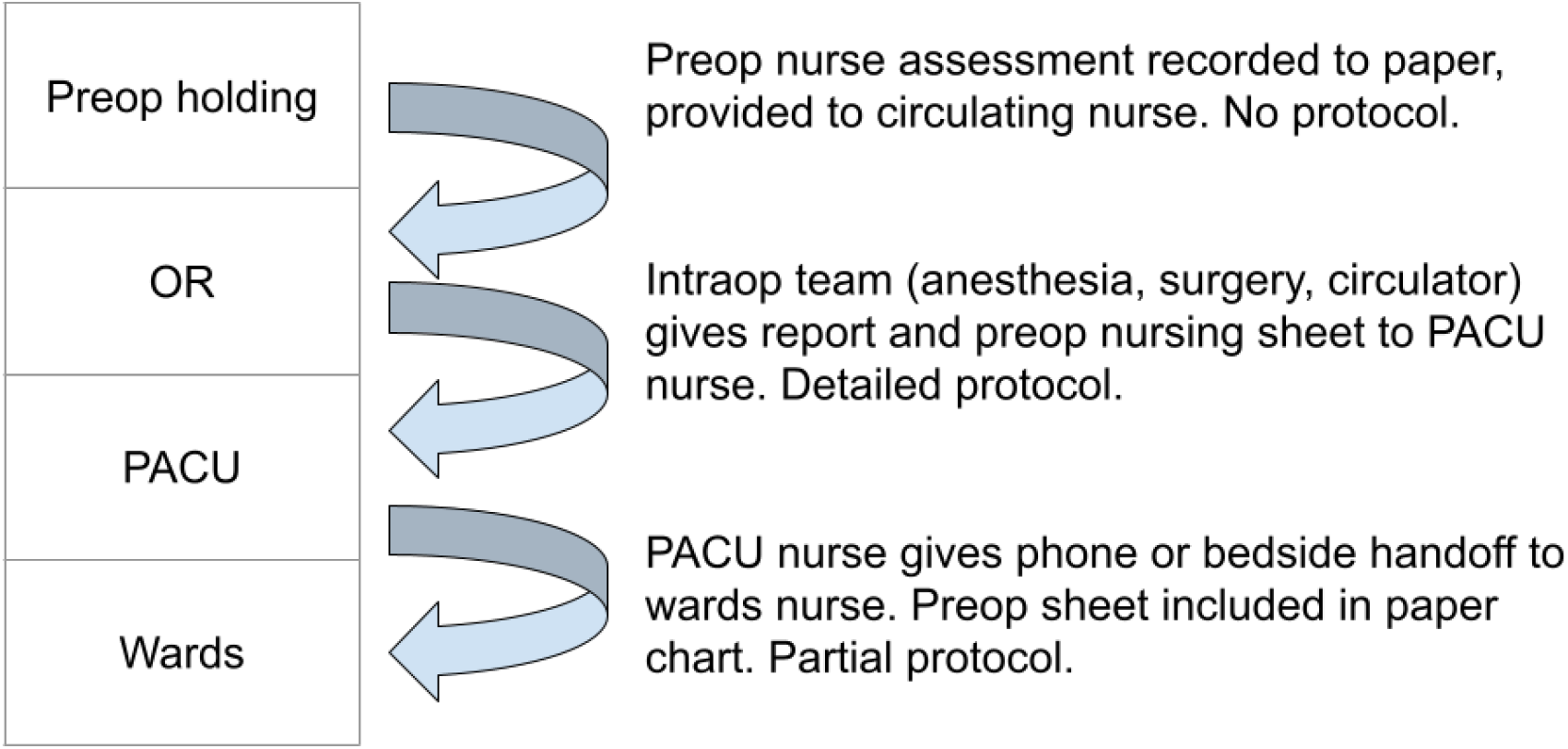
Illustration of perioperative handoff stages

Once a patient is deemed ready to leave the PACU, handoff is given from the PACU nurse to the ward nurse either at the bedside (high dependency unit) or by phone call (other units). A guideline addresses the handoff between PACU and the wards (Appendix S1). Postoperative patients are jointly managed by fellows, resident physicians, nurse practitioners, and the attending surgeon. The nurse practitioner or resident implementing ward care is not directly involved in the surgery. For clarity’s sake, we will refer to that resident or nurse practitioner as the *ward physician*.

### Data collection

Eligible patients were identified from the operating room status board between 9am and 5pm on weekdays, and all eligible patients were attempted for observation. A researcher approached PACU nurses before patient arrival for consent to shadow their handoff activities. The participants were observed, and unstructured notes on the OR-PACU handoff process were taken. For patients where the OR-PACU handoff was missed because of insufficient time to identify the patient’s bed assignment, the nurse was approached for consent to shadow the PACU-ward handoff. We asked the PACU nurse to alert us when a handoff was imminent. During handoff observation, we checked for completeness of information along the standard SBAR format, [27] roughly following the same content as prior PACU-specific handoff guides.[40] This (ref 40) checklist was included as a pilot for future studies related to our institutional context, not as primary data collection; we don’t report the resulting scores here. We also assessed if the handoff-giver seemed able to answer the receiver’s questions.

Recruitment for semi-structured interviews occurred during the observation sessions. Our interview questions were focused on handoff communication, patient assessment, physician communication, and potential roles for AI. Interviews were conducted over phone or voice- application (interview guides in Appendix S2). Interviews were recorded and transcribed verbatim; after selection of example quotations, stop words and verbal pauses were removed.

### Analysis

Interviews and observation notes were double-coded by two researchers (King and Shambe) using a mixed inductive-deductive thematic analysis approach.[41] After each interview, the transcript was reviewed, and topical statements identified. Each statement was coded as relevant to OR-PACU, PACU-ward, or both handoffs. Open codes and subthemes were analyzed within handoff type with an inductive hierarchy and revisited to form into major themes. Statements were also coded for relevance to major study questions (Appendix S3), forming the deductive part of the analysis. Codes were compared between coders, and disagreements discussed until consensus. A third researcher (Abraham) was planned to resolve disagreements, but this was not necessary. The OR-PACU and PACU-ward coding trees were compared for similar themes that could be coalesced. Themes were reviewed and revised with Prof. Abraham, and statements not matching a theme reviewed for additional subthemes. After construction of the coding tree, statements were checked to validate their applicability to the higher-level themes. After 10 interviews, a first round of coding was done, and saturation assessed and found to be likely as most topics were addressed by multiple participants; no new topics were found to emerge after the 11^th^ interview.

A consolidated criteria for reporting qualitative research (COREQ)checklist [42] is included in appendix S4 as a qualitative research reporting framework along with some additional methods details.

## RESULTS

27 OR to PACU handoffs and 31 PACU to ward handoffs (partially overlapping patients) were observed. 11 total interviews were conducted: 7 PACU nurses and 4 ward nurses. Table 1 displays the 4 major themes in our findings, subthemes, and exemplar quotations of each subtheme: (1) Nurse understanding of patient condition guides care decision; (2) Handoffs are important to nurse situational awareness; problem focus and information transfer may be improved by AI; (3) AI may augment nurse care decision making and team communication; and (4) user experience and information overload are likely barriers to using AI during handoffs. These themes had substantial interactions, and with each sub-theme we have noted closely related subthemes in square brackets. The relevance to OR-PACU, PACU-ward, or both handoffs and number discussing are located before the exemplar quote.

**Table 1:**
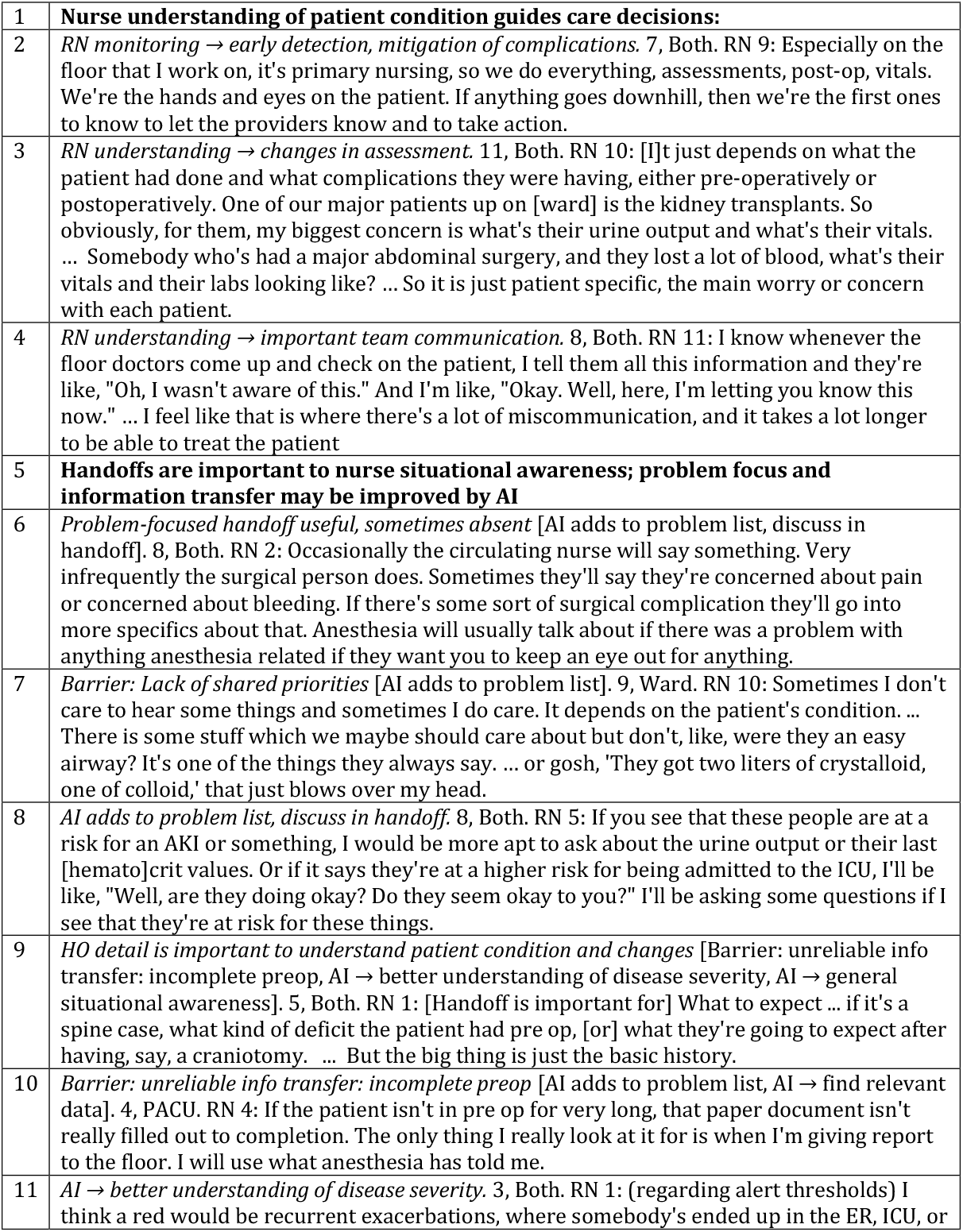
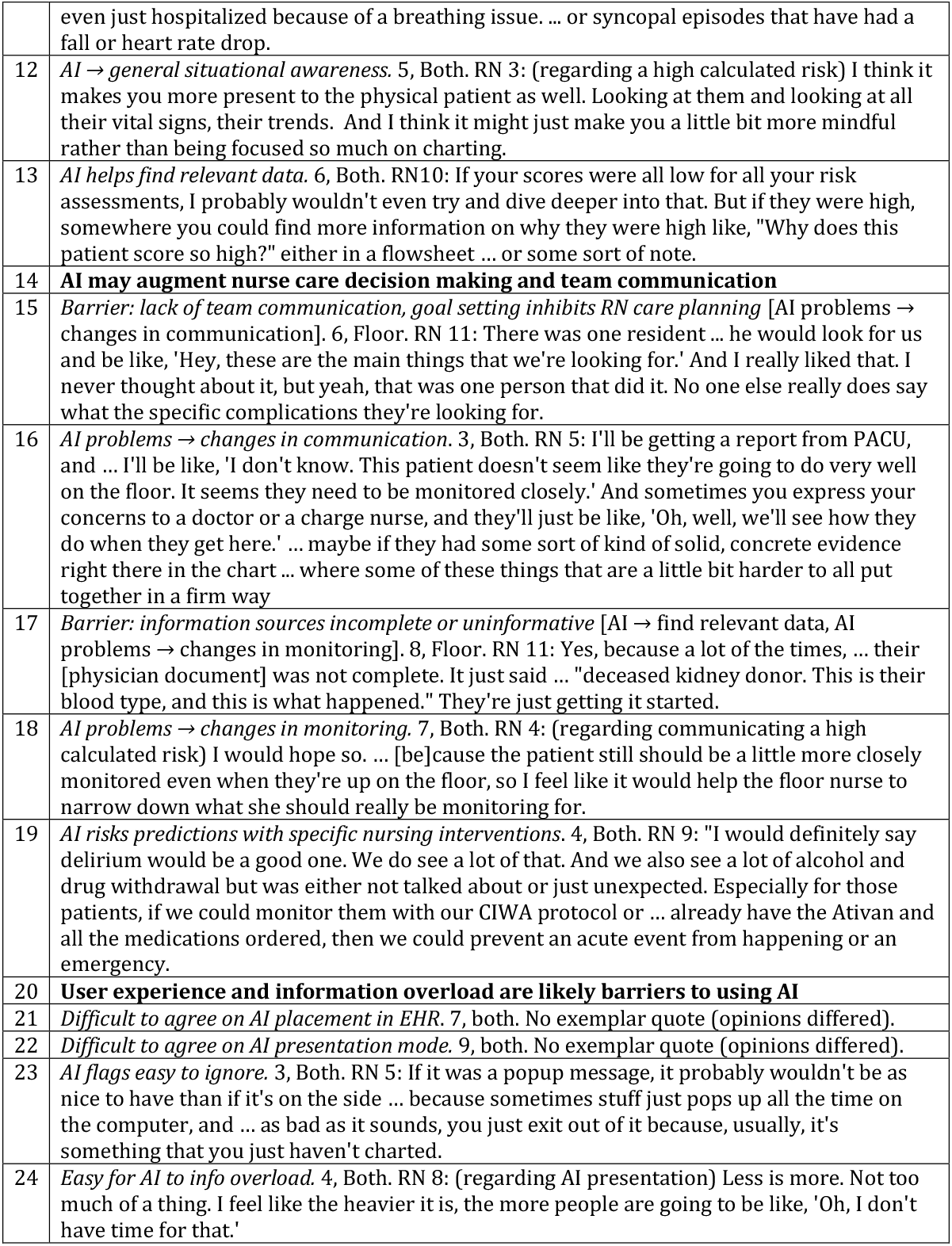
Interview themes, subthemes, and supporting quotations. Subtheme format: theme statement [linked subtheme]. # discussing, relevant to ward vs PACU vs both. Quote.

### Nurse understanding of patient condition guides care decisions

Participants stressed that their bedside presence allowed rapid detection and hopefully mitigation of complications. They universally agreed that their understanding of the issues facing a patient modified what signs and symptoms they were alert for (Table 1 line 3), what issues they communicated to the PACU or ward physician (Table 1 line 4), and what treatments they recommended. Several participants stated that although almost all treatment changes required a team discussion, their recommendations were likely to be considered or acted on.

### Handoffs are important to nurse situational awareness; problem focus and information transfer may be improved by AI

Participants agreed that problem-focused handoffs with anticipatory guidance were extremely useful, but that many topics in handoff were not relevant or recited data without context (Table 1 line 6). Our observations matched this claim: when a problem or complication was under active management, it was likely to be explicitly discussed, but only a fraction of handoffs explicitly discussed potential problems. Closely related to this concern was a lack of shared priorities between the handoff giver and receiver. It was frequent for participants to describe receiving handoffs focusing on details they found to be irrelevant or unintelligible or to not value topics on which their counterparty asked questions (Table 1 line 7). Almost all participants agreed that if AI identified a patient at high risk for a complication, that this topic would be prioritized for discussion at handoff, and that those receiving handoff would ask follow-up questions regarding patient state and the current plan (Table 1 line 8).

Participants stressed that accurate handoff was a critical way to learn about the patient’s state, expectations for recovery, and needs especially in the high-turnover environment of PACU (Table 1 line 9). However, they acknowledged barriers where the documentation they relied on was incomplete (Table 1 line 10), the handoff-giver did not know the relevant information, or they did not understand what needed to be conveyed. Several participants commented on how AI risk prediction at handoff might mitigate mismatch between handoff givers and receivers. First, a high calculated risk could alert them that a known comorbidity was more severe than they expected (Table 1 line 11). Second, awareness that a patient was overall high-risk would prompt nurses to closely review all available data and prioritize careful patient evaluation (Table 1 line 12). Third, automatic identification of EHR data elements which increased the patients’ risk could mitigate data omissions, especially if that data was in an unusual location (Table 1 line 13). Although several participants gave examples of how they might relate data given at handoff to specific AI-identified problems (ameliorating the laundry-list type handoff of Table 1 line 7), none explicitly identified using the AI-identified problems to organize data.

### AI may augment nurse care decision making and team communication

PACU handoff is a critical time for establishing joint plans and physician communication needs. We also noted this in our direct observations; surgery and anesthesiology physicians made requests for notification of symptom or physical status changes and occasionally gave explicit if-then anticipatory guidance. However, ward participants noted that physicians only rarely proactively contacted them, leaving nurses to deduce what issues required communication or nursing action (Table 1 line 15). In our observations, we noted the absence of ward physicians in the overwhelming majority of handoffs. Some participants noted that AI could help target post-handoff nurse-physician communication in two ways. First, if a patient had been identified as high risk, the barrier to contacting the physicians to discuss that topic would be lowered (Table 1 line 16). Second, the nurse’s view of patient risk might be difficult to communicate, and AI-based pattern matching would make this more concrete and easier to request physicians act on or personally evaluate (Table 1 line 16).

Participants noted incomplete physician documentation and other EHR information negatively affected their independent assessment of the patient (table 1 line 17). AI identification of *alternative* key data would then be valuable. Additionally, AI-identified risks for adverse events would allow the nurse to better target their assessment and monitoring independent of any effect on handoff (Table 1 line 18). Participants noted that AI identified elevated risks could allow them to target *interventions* within their scope of practice, such as fall prevention, delirium prevention, and pneumonia prevention (Table 1 line 19). Multiple participants endorsed the desire for more accurate prediction of patients likely to require higher nursing workload or ICU transfer, which they could use to allocate their resources.

### User experience and information overload are likely barriers to using AI

Participants identified several barriers for nursing use of AI, largely centered around the user experience and potential for excessive information volume. First, because of the large number of different methods for accomplishing most tasks in Epic, participants did not recommend the same locations for finding AI risk prediction. Second, preferred visualizations also differed, with participants variously endorsing absolute risk estimates, relative risks, simplified high-medium-low risk flags, and plots. Several participants noted that existing clinical decision support and alerts already generate alarm fatigue, and that additional flags would likely be ignored (Table 1 line 23). Finally, participants noted the potential for information overload with more complex outputs (Table 1 line 24).

### Other barriers to perioperative handoffs

Participants noted many other barriers and several facilitators to perioperative handoffs and patient care. These other findings, mostly reflecting local practices, are documented in appendix S5 as they are outside the scope of the goals of this study.

## DISCUSSION AND CONCLUSION

Our interviews highlighted the importance of team communication and anticipatory guidance at and around postoperative handoffs for nurses to optimize patient care. The data gave consistent answers to our knowledge-gap questions: (1) participants believed that AI which identified patients at elevated risk would lead to focused handoff communication and physician-nurse team communication on those topics, increasing anticipatory guidance and situational awareness. (2) Participants believed that well-functioning AI risk assessment would lead to activating nurse-driven interventions. To accomplish this, participants desired both overall measures of acuity and estimation of a broad collection of risks. (3) While our participants were enthusiastic for AI identification of relevant information in the EHR, they also acknowledged barriers surrounding the user-experience of adding AI to their workflows and the potential for information overload.

Our work contrasts with much of the development of AI EHR support for nurses, [31] which largely focuses on medication documentation, medication administration, and very simple rule-based systems to identify specific nursing needs. Our work also highlights the need for handoff communication to adapt to the patient’s condition, contrasting with the dominant theme of the literature for improving handoffs: standardized communication and checklists.[43] Similar themes have been found in several small studies from other nursing contexts. Home care nurses in a prior study expressed a similar use case for AI to modify the intensity of their services but did not discuss its role in transitions of care.[29] User-design work for EHR-integrated shift-change handoff support had similar ideas, arriving at a design which blended data and predictive risks.[32, 38] Although their work stemmed from interactions with nurses and nursing students, they do not describe the methods to arrive at these conclusions or supporting beliefs by nurses. A prototype system for shift-change in the neonatal intensive care unit focused on summarizing data in natural language, and included expert decision rules as a minor component.[33] Our findings can also be viewed as similar to prior work with dashboards intended to detect change in patient status which lack explicit AI predictions.[31] In our work on OR to ICU handoffs, [39] participants endorsed similar desires to integrate AI into summaries of patient data like laboratory results and vital signs and the need to focus on actionability. In contrast to ICU participants, our participants felt that AI augmentation of handoff topics could be useful, AI assessment of risks for physician communication would be valuable, and that AI could assist their selection of necessary patient assessment steps. Taken together, our findings and these prior studies suggest that AI can support nurses in their more general cognitive tasks, and that future AI design efforts should (1) target critical moments of evaluation like shift change and handoff and (2) incorporate estimates of severity and influential data outside narrow “nursing related” problems.

### Limitations

Our study drew participants from a single center, which limits the range of experiences. The ward nurses worked in a small number of units, limiting the diversity of views. The setting was an academic medical center, so the views may not reflect the experiences of those in private practice. Our interview was semi-structured (but nearly structured), and participants were informed on the nature of our study. They may have endorsed ideas to be agreeable, but participants seemed to feel free to disagree.

## Supporting information

Combined supplement

## Data Availability

Data are not publicly shared. To encourage frank discussion of their workplace and patient care, participants were offered that their transcripts would not be shared.

## Acknowledgment

Several Barnes-Jewish Hospital perioperative nursing leaders helped encourage participation in the study; we would like to thank Christa Overmann, John Schweitzer, and Stephanie Spickerman, as well as all our participants.

## Author Contribution Statement

CRK: Conceptualization, Methodology, Formal analysis, Investigation, Data Curation, Writing - Original Draft, Writing - Review & Editing, Project administration, Funding acquisition

JA: Conceptualization, Methodology, Formal analysis, Writing - Review & Editing, Funding acquisition

AS: Formal analysis, Investigation, Data Curation, Writing - Review & Editing

## Data sharing statement

Data cannot be shared for ethical/privacy reasons. The data underlying this article cannot be shared publicly due to identified discussions of provider behavior and patient stories. Participants were offered privacy to allow them to openly share concerns about their workplace. The data will be shared on reasonable request to the corresponding author and IRB approval.

## REFERENCES

1. Keebler, J. R., Lazzara, E. H., Patzer, B. S., Palmer, E. M., Plummer, J. P., Smith, D. C., … Riss, R. (2016). Meta-Analyses of the Effects of Standardized Handoff Protocols on Patient, Provider, and Organizational Outcomes. Human Factors, 58(8), 1187–1205. https://doi.org/10.1177/0018720816672309

2. Solet, D. J., Norvell, J. M., Rutan, G. H., & Frankel, R. M. (2005). Lost in translation: challenges and opportunities in physician-to-physician communication during patient handoffs. Academic Medicine, 80(12), 1094–1099. https://doi.org/10/br5nvk

3. Segall, N., Bonifacio, A. S., Schroeder, R. A., Barbeito, A., Rogers, D., Thornlow, D. K., … Inquiry, O. behalf of the D. V. P. S. C. of. (2012). Can We Make Postoperative Patient Handovers Safer? A Systematic Review of the Literature. Anesthesia & Analgesia, 115(1), 102. https://doi.org/10/gmf4bm

4. Nagpal, K., Arora, S., Abboudi, M., Vats, A., Wong, H. W., Manchanda, C., … Moorthy, K. (2010). Postoperative Handover: Problems, Pitfalls, and Prevention of Error. Annals of Surgery, 252(1), 171–176. https://doi.org/10/chw43w

5. Nagpal, K., Arora, S., Vats, A., Wong, H. W., Sevdalis, N., Vincent, C., & Moorthy, K. (2012). Failures in communication and information transfer across the surgical care pathway: interview study. BMJ Quality & Safety, 21(10), 843–849. https://doi.org/10.1136/bmjqs-2012-000886

6. Hughes, H. K., Serwint, J. R., O’Toole, J. K., Spector, N. D., & Ngo, T. L. (2019). I-PASS Adherence and Implications for Future Handoff Training. Journal of Graduate Medical Education, 11(3), 301–306. https://doi.org/10/gghzph

7. Hughes, T. M., Dossett, L. A., Hawley, S. T., & Telem, D. A. (2019). Recognizing Heuristics and Bias in Clinical Decision-Making. Annals of Surgery. https://doi.org/10/gghzpg

8. Wheeler, D. S., Sheets, A. M., & Ryckman, F. C. (2018). Improving transitions of care between the operating room and intensive care unit. Translational Pediatrics, 7(4), 299– 307. https://doi.org/10/gmf4cf

9. Wright, M. C., & Endsley, M. R. (2008). Building Shared Situation Awareness in Healthcare Settings. In Improving Healthcare Team Communication. CRC Press.

10. Bergman, A. A., Flanagan, M. E., Ebright, P. R., O’Brien, C. M., & Frankel, R. M. (2016). “Mr Smith’s been our problem child today…”: anticipatory management communication (AMC) in VA end-of-shift medicine and nursing handoffs. BMJ Quality & Safety, 25(2), 84– 91. https://doi.org/10/f796tk

11. Horwitz, L. I., Moin, T., Krumholz, H. M., Wang, L., & Bradley, E. H. (2008). Consequences of Inadequate Sign-out for Patient Care. Archives of Internal Medicine, 168(16), 1755– 1760. https://doi.org/10/ctnpvr

12. Starmer, A. J., Schnock, K. O., Lyons, A., Hehn, R. S., Graham, D. A., Keohane, C., & Landrigan, C. P. (2017). Effects of the I-PASS Nursing Handoff Bundle on communication quality and workflow. BMJ Quality & Safety, 26(12), 949–957. https://doi.org/10/fphf

13. Sheth, S., McCarthy, E., Kipps, A. K., Wood, M., Roth, S. J., Sharek, P. J., & Shin, A. Y. (2016). Changes in Efficiency and Safety Culture After Integration of an I-PASS–Supported Handoff Process. Pediatrics, 137(2), e20150166. https://doi.org/10/f8cbcq

14. Shahid, S., & Thomas, S. (2018). Situation, Background, Assessment, Recommendation (SBAR) Communication Tool for Handoff in Health Care – A Narrative Review. Safety in Health, 4(1), 7. https://doi.org/10/fspc

15. Philibert, I. (2009). Use of strategies from high-reliability organisations to the patient hand-off by resident physicians: practical implications. BMJ Quality & Safety, 18(4), 261–266. https://doi.org/10.1136/qshc.2008.031609

16. Zjadewicz, K., Deemer, K. S., Coulthard, J., Doig, C. J., & Boiteau, P. J. (2018). Identifying What Is Known About Improving Operating Room to Intensive Care Handovers: A Scoping Review. American Journal of Medical Quality, 33(5), 540–548. https://doi.org/10/gmf4bh

17. McFarlane, A. (2018). The impact of standardised perioperative handover protocols: Journal of Perioperative Practice. https://doi.org/10/gh3427

18. Staggers, N., & Blaz, J. W. (2013). Research on nursing handoffs for medical and surgical settings: an integrative review. Journal of Advanced Nursing, 69(2), 247–262. https://doi.org/10/f4m595

19. Street, M., Phillips, N. M., Haesler, E., & Kent, B. (2018). Refining nursing assessment and management with a new postanaesthetic care discharge tool to minimize surgical patient risk. Journal of Advanced Nursing, 74(11), 2566–2576. https://doi.org/10/ggchqp

20. Street, M., Phillips, N. M., Mohebbi, M., & Kent, B. (2017). Effect of a newly designed observation, response and discharge chart in the Post Anaesthesia Care Unit on patient outcomes: a quasi-expermental study in Australia. BMJ Open, 7(12). https://doi.org/10/gcnr2x

21. Siddiqui, N., Arzola, C., Iqbal, M., Sritharan, K., Guerina, L., Chung, F., & Friedman, Z. (2012). Deficits in information transfer between anaesthesiologist and postanaesthesia care unit staff: an analysis of patient handover. European Journal of Anaesthesiology, 29(9), 438–445. https://doi.org/10.1097/EJA.0b013e3283543e43

22. Milby, A., Böhmer, A., Gerbershagen, M. U., Joppich, R., & Wappler, F. (2014). Quality of post-operative patient handover in the post-anaesthesia care unit: a prospective analysis. Acta Anaesthesiologica Scandinavica, 58(2), 192–197. https://doi.org/10.1111/aas.12249

23. Halladay, M. L., Thompson, J. A., & Vacchiano, C. A. (2018). Enhancing the Quality of the Anesthesia to Postanesthesia Care Unit Patient Transfer Through Use of an Electronic Medical Record–Based Handoff Tool. Journal of PeriAnesthesia Nursing. https://doi.org/10.1016/j.jopan.2018.09.002

24. Pucher, P. H., Johnston, M. J., Aggarwal, R., Arora, S., & Darzi, A. (2015). Effectiveness of interventions to improve patient handover in surgery: A systematic review. Surgery, 158(1), 85–95. https://doi.org/10.1016/j.surg.2015.02.017

25. Flemming, D., & Hübner, U. (2013). How to improve change of shift handovers and collaborative grounding and what role does the electronic patient record system play? Results of a systematic literature review. International Journal of Medical Informatics, 82(7), 580–592. https://doi.org/10/f2w36d

26. Staggers, N., Clark, L., Blaz, J. W., & Kapsandoy, S. (2011). Why patient summaries in electronic health records do not provide the cognitive support necessary for nurses’ handoffs on medical and surgical units: insights from interviews and observations. Health Informatics Journal, 17(3), 209–223. https://doi.org/10/cs8qht

27. Staggers, N., Clark, L., Blaz, J. W., & Kapsandoy, S. (2012). Nurses’ information management and use of electronic tools during acute care handoffs. Western Journal of Nursing Research, 34(2), 153–173. https://doi.org/10/fsnqp6

28. Waller, R. G., Wright, M. C., Segall, N., Nesbitt, P., Reese, T., Borbolla, D., & Del Fiol, G. (2019). Novel displays of patient information in critical care settings: a systematic review. Journal of the American Medical Informatics Association, 26(5), 479–489. https://doi.org/10.1093/jamia/ocy193

29. Dowding, D. W., Russell, D., Onorato, N., & Merrill, J. A. (2017). Technology Solutions to Support Care Continuity in Home Care: A Focus Group Study. Journal for Healthcare Quality, 1. https://doi.org/10/gmf4b7

30. Dowding, D., Merrill, J. A., Barrón, Y., Onorato, N., Jonas, K., & Russell, D. (2019). Usability Evaluation of a Dashboard for Home Care Nurses. CIN: Computers, Informatics, Nursing, 37(1), 11. https://doi.org/10/gmf4b6

31. Dunn Lopez, K., Gephart, S. M., Raszewski, R., Sousa, V., Shehorn, L. E., & Abraham, J. (2017). Integrative review of clinical decision support for registered nurses in acute care settings. Journal of the American Medical Informatics Association, 24(2), 441–450. https://doi.org/10/gddxrt

32. Forbes, A., Surdeanu, M., Jansen, P., & Carrington, J. (2013). Transmitting Narrative: An Interactive Shift-Summarization Tool for Improving Nurse Communication. In Proceedings of the 3rd IEEE Workshop on Interactive Visual Text Analytics. Retrieved from http://clulab.cs.arizona.edu/papers/textvis2013.pdf

33. Hunter, J., Freer, Y., Gatt, A., Reiter, E., Sripada, S., & Sykes, C. (2012). Automatic generation of natural language nursing shift summaries in neonatal intensive care: BT-Nurse. Artificial Intelligence in Medicine, 56(3), 157–172. https://doi.org/10/f4j6j9

34. Xue, B., Li, D., Lu, C., King, C. R., Wildes, T., Avidan, M. S., … Abraham, J. (2021). Use of Machine Learning to Develop and Evaluate Models Using Preoperative and Intraoperative Data to Identify Risks of Postoperative Complications. JAMA network open, 4(3), e212240. https://doi.org/10/gjtj6s

35. Ting, J. J., & Garnett, A. (2021). E-Health Decision Support Technologies in the Prevention and Management of Pressure Ulcers: A Systematic Review. Computers, informatics, nursing: CIN. https://doi.org/10/gmf4bz

36. Hu, Y.-H., Lee, Y.-L., Kang, M.-F., & Lee, P.-J. (2020). Constructing Inpatient Pressure Injury Prediction Models Using Machine Learning Techniques. Computers, informatics, nursing: CIN, 38(8), 415–423. https://doi.org/10/gh6fbv

37. Lindberg, D. S., Prosperi, M., Bjarnadottir, R. I., Thomas, J., Crane, M., Chen, Z., … Lucero, R. J. (2020). Identification of important factors in an inpatient fall risk prediction model to improve the quality of care using EHR and electronic administrative data: A machine-learning approach. International Journal of Medical Informatics, 143, 104272. https://doi.org/10/gmf4b4

38. Chetta, A., Carrington, J. M., & Forbes, A. G. (2015). Augmenting EHR interfaces for enhanced nurse communication and decision making. In Proceedings of the 2015 Workshop on Visual Analytics in Healthcare (pp. 1–6). New York, NY, USA: Association for Computing Machinery. https://doi.org/10/gg5fxp

39. Abraham, J., King, C. R., & Meng, A. (2021). Ascertaining Design Requirements for Postoperative Care Transition Interventions. Applied Clinical Informatics, 12(1), 107–115. https://doi.org/10/gmf38t

40. Weinger, M. B., Slagle, J. M., Kuntz, A. H., Schildcrout, J. S., Banerjee, A., Mercaldo, N. D., … France, D. J. (2015). A Multimodal Intervention Improves Postanesthesia Care Unit Handovers. Anesthesia and Analgesia, 121(4), 957–971. https://doi.org/10/ggj6zc

41. Braun, V., Clarke, V., Hayfield, N., & Terry, G. (2019). Thematic Analysis. In P. Liamputtong (Ed.), Handbook of Research Methods in Health Social Sciences (pp. 843– 860). Singapore: Springer Singapore. https://doi.org/10.1007/978-981-10-5251-4_103

42. Tong, A., Sainsbury, P., & Craig, J. (2007). Consolidated criteria for reporting qualitative research (COREQ): a 32-item checklist for interviews and focus groups. International Journal for Quality in Health Care, 19(6), 349–357. https://doi.org/10/fm8b7v

43. Hilligoss, B., & Moffatt-Bruce, S. D. (2014). The limits of checklists: handoff and narrative thinking. BMJ Quality & Safety, 23(7), 528–533. https://doi.org/10/f572m2

