## Supplementary material for "Potential uses of AI for perioperative nursing handoffs: a qualitative study": Combined supplement

Appendix S1: Handoff artifacts and protocols in use.

Figure S1.1: Preop nursing assessment sheet. This paper form is begun by an RN in the preop holding area while interviewing a patient, filling out the top four sections. The form is placed in the paper chart, where it accompanies the patient to the OR, PACU, and wards. The PACU RN fills out the “Intraop” and “Postop” sections. We found that this form was used in every OR-PACU handoff and almost every PACU-floor handoff.

### CHART HAND OFF WORKSHEET

**PT INFO**

Name:

Case Number:

NKDA Allergy

**PROCEDURE:**

**HISTORY**

☐ HTN ☐ Asthma
☐ COPD ☐ Seizures
☐ Chronic pain ☐ GERD
☐ Cardiac history ☐ OSA - CPAP
☐ Parkinsons

Other:

DM Type 1 home insulin pump

DM Type 2

Fall Risk: ☐ yes ☐ no

Translator:

Isolation:

Implantable Device:

Outpatient: ☐ yes ☐ no

Family Member:

Belongings:

Phone number:

**PREOP**

VS:

Mental Status:

Skin Integrity:

Betablockers: ☐ yes ☐ no

Home Opioids: ☐ yes ☐ no

HCG Results: ☐ Positive ☐ Negative

Accucheck:

**REGIONAL**

PREOP
☐ Regional Block
☐ Spinal
☐ Epidural

POTENTIAL POST OP BLOCK
☐ Yes → Call Regional Anesthesia
☐ No → Reason: ____________________
☐ Maybe → Call Regional Anesthesia
Dermatome ____________________

Post-op Block ______________ Time: ________

RAS Nurse 314-362-3907

Regional Anesthesia phone: ____________________

**INTRAOP**

| | | |
|---|---|---|
| General | MAC | Spinal |
| ET | LMA | Epidural |
| Airway: | NP | ORAL |

Insulin:

| | |
|---|---|
| Versed | Zofran |
| Fentanyl | Pepcid |
| Dilaudid | Benadryl |
| Toradol | Decadron |
| Antibiotic | |

Crystalloid
Colloid
EBL
U/O
Blood products

Complications

Skin

Relaxed / Reversed

Family Last Updated:

Positioning

**POSTOP**

IV Access

Fluids/Gtts

Skin Breakdown

Assessment:

Surgical Wound:

Concerns:

DRAINS
JP
CT
HV
Lumbar
Ventric

COMPASS ORDERS
CXR
Hip/Knee/Pelvis
PCA

LABS
CBC
BMP
Coags
Trops
Glucose
Other:

**Misc/To Do:**

Surgical MD Phone Number:

Sign Out Time:

Boarding Time:

Destination:



**PLACE IN FRONT OF CHART**

Figure S1.2: OR to PACU handoff protocol. A laminated printout of this document hangs by every PACU bay.

### Handoff Critical Elements

#### Universal Handoff Process

- Receiving team begins critical monitor hookup
  - Report cannot start until hookup is complete
- Team members introduce themselves

#### Circulator

- Patient identification (confirm placement of wristband)
- Isolation type
- Position of patient intra-operatively
- Packing/retained items
- Family information/last update
- Belongings/valuables
- Special equipment
- Key intraoperative events
- "The thing I am most concerned about is..."
- "What other questions do you have for me?"

#### Surgical/Procedure

- Indication for surgery/procedure
- Baseline physical exam – neuro, demeanor, pertinent positives
- Expected post-op exam findings (known neurologic deficits, expected pulse check, etc)
- Drain/tubes/packing/ intentionally retained surgical items (location, number, types and labeled)
- Dressing/wound
- Complications
- Labs
- Imaging (including providers needing to review prior to discharge from PACU)
- Ability for alternate pain management – peripheral block, epidural, TAP
- Diet (including PO and per tube medications)
- Special considerations (positioning, hemodynamic/ flap check parameters if necessary)
- Contact information for service
- Patient disposition plan (home, floor, ICU)
- "The thing I am most concerned about is..."
- "What other questions do you have for me?"

#### Anesthesiology

- PMH/PSH
- Allergies
- Meds (specify which were taken prior to surgery relate to HTN, DM, Parkinson's, chronic pain)
- Baseline vitals; height, weight, pain
- Baseline labs of significanc[illegible]
- Airway
- Lines
- Procedures (blocks, spinals
- Fluids (EBL and blood or blood products)
- Paralytic status
- Labs
- Meds (vasopressors, last dose of antibiotics, other intraoperative medications and infusions)
- Key events (hemodynamic stability)
- Pain management for this patient will consist of...
- Code status (including any conversations about temporary suspensions of DNR or DNI)
- Orders to follow up on (labs blood glucose, CXR for ___
- "The thing I am most concerned about is..."
- "What other questions do you have for me?"

Figure S1.3: PACU to Ward handoff protocol. This document is the reverse side of Figure S1.2.

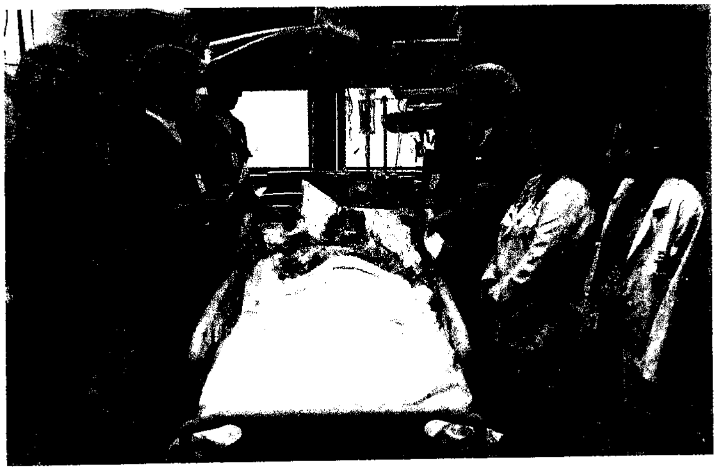

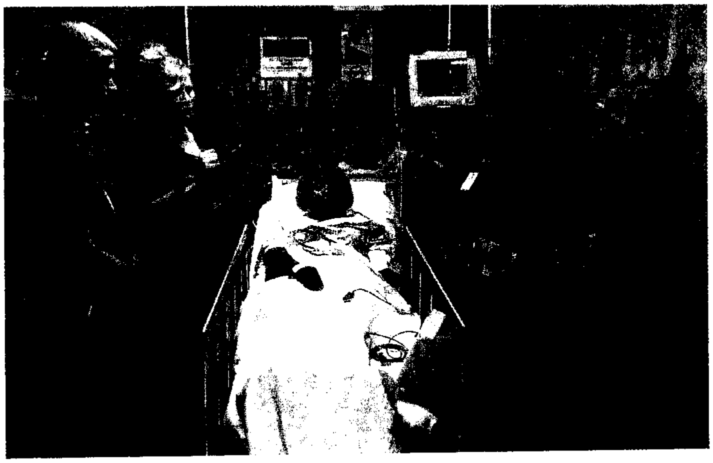

#### Patients Being Discharged From PACU

- Identification of patient and procedure
- Important past medical history
- Any complications intraoperative
- Any complications postoperative
- I and O's
- Labs/images results or pending
- Medications given and resolutions
- Regional blocks
- OSA risk
- Patient placement
- Telemetry ordered

PACU Nurse Interview Guide

Intro statement – review consent and study

Thank you so much for participating. To review the study consent document we went over, this will be an interview about handoffs in the PACU and how predictive analytics might fit into your workflow. This is an IRB approved study, and we will be recording the conversation and transcribing it. You can withdraw at any time, and I haven't offered your any compensation. Does that match your understanding?

Intro statement – topic

To set up context, do you primarily work in PACU or do you also work in preop or other areas? Do you work only at BJH or other hospitals as well?

1. What would you say the contribution of the PACU is to preventing postop complications?

I understand that when a patient comes from the OR, after they are hooked up to the monitor, and then the circulator, surgeon, and anesthesia provider give report.

2. How often is the handoff protocol fully followed? Are there any major omissions that happen?
3. Is there information that you routinely need to look for in Epic after handoff?
4. Is there information you wish was easier to get to?
5. Do you talk about patient specific risks of complications at handoff?

6. How does your monitoring and evaluation vary from patient to patient in PACU?
    a. Do you spend more time with patients or change what you're looking for based on their comorbidities or surgery?
    b. Does your communication with the surgery service or PACU anesthesia team vary based on comorbidities or surgery?

I'd like to ask a few questions about the communication to the floor. My understanding is that after the patient has met some PACU goals and anesthesia has signed them out, you call the floor nurse or transport with them and do a face to face handoff.

7. Is there a protocol for what needs to be communicated to the floor nurse?
8. Do you give any information to the floor provider (resident or NP)?
9. Are there questions the postop nurse frequently asks?
10. Do you talk about patient-specific risks of complications?
11. What do you think that we could do that would most improve the transfer of important information to the postop wards team?

I'd like to shift focus a bit and talk about the potential role of predictive analytics for PACU nurses. Epic and other investigators are looking into displaying machine learning predicted risks of common adverse events like pneumonia, delirium, and AKI into everyone's workflow. For context, you can assume that we are really only interested in more complicated cases with higher risk patients or surgeries.

12. Are there any specific adverse events where these calculated risks that would be useful for you to know?

a. if it was present at entry to the PACU, would it affect what you looked for in handoff?
b. would it affect communication with anesth / surgery while in PACU
c. do you think knowing elevated risks like these might affect what you're looking for with the patient?
d. If this sort of information was integrated to Epic or included as a print-out with the preop nursing sheet, would you refer to it in your handoff to the floor?

13. Would it be useful to you to have more general risks like length of stay, ICU admission, and death? How so?
14. Let's say that something like ICU admission risk was going to be added. Can you think where in your workflow it might be reasonable to look at?
15. Again assuming that these were going to show up, are there thresholds in any of these risks that make them relevant? Would you like to see the numbers, a graph, or a simplified presentation (like low, medium, high)?
    a. Would it be helpful to include some comparisons like an "average patient", patients getting this surgery?
    b. Would it be helpful to know how the risks changed during the OR useful?
16. A feature that's also been suggested is the link between risk factors and predictive analytic risks. For example, that the risk of ICU admission was high because of anemia and low albumin. This can also be shown quantitatively (e.g. the risk of ICU admission increased by 3% due to the low hemoglobin). Do you think those would be useful?
    a. What about (LOS is high) because (pneumonia risk is high)

Ward Nurse Interview Guide

Intro statement – review consent and study

Thank you so much for participating. To review the study consent document we went over, this will be an interview about handoffs from the PACU and how predictive analytics might fit into your workflow. This is an IRB approved study, and we will be recording the conversation and transcribing it. You can withdraw at any time, and I haven't offered your any compensation. Does that match your understanding?

1. What units / areas do you normally work in?
2. I have a big picture question to get started thinking about the topic: what would you say the contribution of the ward nurse is to preventing postop complications?
3. Tell me briefly about what happens when a patient comes from the PACU. (if not specifically addressed)
    a. Is there a specific handoff protocol that is followed? What are the elements of it? What fraction of the time is it completely followed?
    b. What specific tasks do you need to accomplish right away when a patient arrives?
4. Tell me about the information you get from handoff with the PACU.
    a. Do you normally get any information from the surgery resident / np?
    b. Do you think this is fairly complete in terms of understanding what happened in the OR and PACU? Is there anything important that is sometimes missed?
5. Is there information that you routinely look for in Epic after handoff?
    a. Do you look at the surgery NP "handoff" document?
    b. Is there information you wish was easier to get to?
    c. Is there any information that you rely on the paper chart for?
6. For immediate postop patients, how does the monitoring vary from patient to patient?
    a. Do you spend more time with patients or change what you're looking for based on their comorbidities or surgery?
7. As far as variation in nursing interventions go, are there some that you have the leeway to decide them on your own, and some that you reach out to the NP / physician for changes to orders? Can you briefly given an example?
8. What communication does the surgical team routinely expect from you?
    a. Other than an emergency or forgotten orders, what would you call the surgery team for?
    b. What do they get in touch with you for?
    c. Do you talk about patient-specific risks?
9. *Skip if > 15 minutes.* If you handoff the patient to another nurse before shift change from time to time is there a specific protocol for nurse-nurse handoffs?
    a. How do you share information in this case? Is there Epic documentation that you look for when taking over from another nurse?

Another topic we'd like to discuss is predictive analytics. Epic and other investigators are looking into displaying machine learning predicted risks of common adverse events like pneumonia, delirium, and AKI into everyone's workflow. For context, you can assume that we are really only interested in more complicated cases with higher risk patients or surgeries.

These would be generated based on information from the preoperative assessment, medical history, medication list, intraoperative data, and PACU events. We want to hear what might be useful to you and ask a few questions about how it might be presented.

10. Are there any specific adverse events where these calculated risks would be useful for you to know? Some common suggestions are ICU admission, death, AKI, respiratory failure, delirium, length of hospital stay, readmission.
    a. Do you think very specific risks (like pneumonia) are more useful, or more general risk like length of stay?
11. Let's say that something like ICU admission risk was going to be added. Can you think where in your workflow it might be reasonable to look at?
12. If some of these risks were going to be shown, would it be useful to have a comparison like an "average patient" or just show the absolute risk?
13. Would you like to see the numbers, a graph, or a simplified presentation (like low, medium, high)? Would the change over the course of the hospitalization or surgery be useful to show?
14. A feature that's also been suggested is the link between risk factors and predicted risks. For example, that the risk of ICU admission was high because the albumin was low. This can be shown qualitatively (like, just a list of risk factors) or quantitatively (e.g. the risk of ICU admission increased by 3% due to the low hemoglobin). Do you think those would be useful?
15. If this sort of information was integrated to Epic would you refer to it in your handoff from PACU? Do you think that you would refer to it in your documentation? What about the list of identified risk factors?

Finally, we'd like to hear any thoughts you have on these issues that we didn't touch on.

16. What do you think that we could do that would most improve the transfer of important information to the postop nursing team?

**Appendix S3: Core study topics used as closed codes**

Description of handoff process

- Process failures, common information difficulties
- Protocol use / awareness
- Anticipatory guidance topics
- Information sought / sought by others

Nurse decision making

- Variations in care
- Communication with team and autonomous decisions

Use of AI

- Willingness to use / characteristics of effective
- Use cases: specific recommendations, physician or handoff communication, other
- Specific risks worth predicting

Presentation of AI

- Preferred presentation
- Workflow placement
- Feature attribution / data seeking

Additional COREQ Checklist Information

Domain 1: Research team and reflexivity

Interviews and recruitment were conducted by CRK and AOS. Direct observations were performed by AOS.

CRK has been an attending anesthesiologist at the facility for the previous 2 years and a trainee at the institution prior to that. He is a KL2 scholar focusing on user-centered implementation of clinical decision support with coursework in qualitative data collection. He was professionally known to most PACU staff but previously known to very few postop staff. Participants were informed that CRK was working on clinical decision support as part of the introduction and consent steps. He has a professional investment in the research question.

AOS is a research assistant and entering medical student at a nearby university, and had no previous contact with the participants. She has prior training in interview-based qualitative data collection and analysis, but no prior exposure to or beliefs about the study topic.

Domain 2: Study design

Methodological orientation and Theory: content analysis
Sampling: Convenience
Method of approach: In the direct observation phase, we explained the study, and requested their informed consent to shadow their handoff process. We approached the PACU or ward nurse for interviews by speaking at unit-wide "huddles", emails to unit email list-serves, posting flyers in unit breakrooms, and one-on-one discussions after direct observation of patient handoffs. After funding became available, potential participants were incentivised with $50 gift cards.
Sample size: 11
Non-participation: exact numbers of approaches for direct observation were not recorded, but the great majority agreed. Conversely, fewer than 1/3 of approached participants agreed to an interview.
Setting of data collection: Observation data was collected in the hospital (workplace). Participants selected interview times, and most participated from their homes; a small number conducted the interview from a break room or empty patient room.
Presence of nonparticipants: Other than the two researchers, no non-participants were present.
Description of sample: 10/11 participants were female; 10/11 participants were white; all participants were working registered nurses. Ages were not collected.
Interview guide: provided in a separate appendix. No additional testing was performed.
Repeat interviews: none
Audio/visual recording: phone or voice-application (e.g. Zoom, Teams), audio recorded, video declined by all participants.
Field notes: Yes
Duration: 30-45 minutes

Data saturation: saturation was discussed between CRK and AOS when stopping interview recruitment.
Transcripts returned: No

Domain 3: analysis and findings
Number of data coders: CRK and AOS coded
Description of the coding tree: subthemes and themes linked in Table 1
Derivation of themes: mixed inductive and deductive. Subthemes and themes reviewed and revised with JA. Open coded data subsequently reviewed for fit with final themes and subthemes.
Software: Excel, R
Participant checking: Participants were not invited for the analysis
Quotations presented: in Table 1
Data and findings consistent: yes
Clarity of major themes: Majority of results presentation
Clarity of minor themes: subthemes discussed

**Appendix S5: Other findings regarding handoff**

Although an OR to PACU protocol is in place, violations of that protocol were fairly common. RN 2: "There is, there is one that's supposed to be following. Very few people actually follow it. Usually it's the newer people who follow it. I mean it's not complicated, so ..." Our direct observations supported several common protocol violations. First, surgery providers typically left before anesthesia providers gave report. It is unclear if any relevant information had already been conveyed to them. Participants noted (and we observed) that occasionally no qualified member of the surgical team accompanied the patient to PACU, resulting in inadequate handoff and necessitating several additional phone calls to establish a plan. RN 4: "a lot of times they at least try to send out a first assist, but there have been a few occasions where certain services, like if they don't have a fellow or resident working with them, they just won't come out and they'll just say call into the room that sometimes can be a gap."

Many of the notes made by the PACU nursing staff are not documented in the EHR. They are recorded on paper, which is not subsequently used. RN 4: "I don't know if it's something that needs to be like documented, in epic more so everybody can see it. 'cause I know sometimes, like we write on that handoff paper, but sometimes that gets misplaced. I don't honestly think the floor really looks at it at all."

Participants noted several barriers for communication coming out of PACU. First, PACU uncommonly directly contacted the ward physician to update them on patient status. RN 3: "I personally don't speak to and I don't believe that any of their other nurses speak to the NPs or or residents from the floor." Additionally, the PACU nursing divides its communication between the PACU anesthesiology team and surgery service, further reducing the communication pathways for the floor providers. RN 3: "The anesthesia team is primarily our source of contact for patients when acute things do happen, they come before this surgical service because they're kind of there being able to manage the patients. ... Any kind of vital sign issue, acute agitation or things like that. ... If it were something that was going to be needing addressed outside of the operative recovery space, that would be more primary [service]."

Although a standard SBAR report is expected (Appendix 1), no participants acknowledged a protocol for PACU to floor handoff. RN 6: "I believe no, not necessarily a direct protocol. There's no hand-off sheet. We kind of use the sheets that we do report with, or at least I do, and I go down that list. But there isn't-- at least in the PACU. there's not a set 'this is everything to go in this exact order,' like we have the hand-off from the OR to us."

Handoffs around shift change are particularly fraught, both because the giving team has lost information and because the receiving team is busy with multiple tasks: RN 7: "I think that sometimes what's missed is we get the patients after the next shift has started and that PACU nurse maybe only had them for a half-hour or so, and so they don't know a ton about them either. And then they'll bring up four PACU patients back to back within like half an hour. And we're not that big, so it takes three or so staff members to get the other patients checked in. And so sometimes I feel like things can be missed that way."

One facilitator of care which participants noted was that they had a low barrier to requesting input from the PACU or ward physician, or to notify them of changes in patient status.  RN 7: "If there's any deviation or if we see a trend of something like the urine going down drastically, we call them. We pretty much call them on some of these patients every hour."
